# The Asia-Pacific body mass index classification and new-onset chronic kidney disease in non-diabetic Japanese adults: A community-based longitudinal study from 1998 to 2023

**DOI:** 10.1101/2024.01.06.23300566

**Authors:** Yukari Okawa, Toshiharu Mitsuhashi, Toshihide Tsuda

## Abstract

**Background/Objectives:** Obesity is a risk factor for chronic kidney disease (CKD) in Asians. The Asia-Pacific body mass index (BMI) classification sets lower obesity cutoffs than the conventional BMI classification for all races, generally reflecting the lower BMIs in Asians. This longitudinal study evaluated the association between BMI, as classified by the Asia-Pacific BMI system, and CKD development in non-diabetic Asian adults.

**Methods:** A population-based longitudinal study (1998–2023) was conducted in non-diabetic Japanese adults (hemoglobin A1c < 6.5%) in Zentsuji City (Kagawa Prefecture, Japan). The generalized gamma model was used to assess the relationship between time-varying BMI categories and CKD development, stratified by sex. CKD was defined as an estimated glomerular filtration rate of <60 mL/min/1.73 m^2^. BMI was calculated as weight (kg) divided by the square of height (m^2^) and categorized per the Asia-Pacific classification as overweight (23.0–24.9 kg/m^2^), obesity class I (25.0–29.9 kg/m^2^), and obesity class II (≥30.0 kg/m^2^).

**Results:** CKD developed in 34.2% of 3,098 men and 34.8% of 4,391 women. The mean follow-up times were 7.41 years for men and 8.25 years for women. During follow-up, the BMI distributions for men were 5.0% underweight, 43.3% normal weight, 25.6% overweight, 24.1% obesity class I, and 2.0% obesity class II; those for women were 7.7%, 50.5%, 20.5%, 18.3%, and 2.9%, respectively. Compared with normal weight, obesity class I was associated with a 6% (95% confidence interval [CI]: 2%–10%) shorter time to CKD onset in men and 5% (95% CI: 2%–7%) in women. In both sexes, obesity class II showed shorter survival times than normal weight by point estimates, although all 95% CIs crossed the null value.

**Conclusions:** Obesity, as classified by the Asia-Pacific BMI system, shortened the time to CKD onset in non-diabetic Asians. The conventional BMI cutoff for obesity (≥30.0 kg/m^2^) may be too high to identify CKD risk in this population. The findings of this study may be useful for public health professionals in designing interventions to prevent CKD.

## Introduction

Obesity is a well-known risk factor for developing chronic kidney disease (CKD)^1^. Obesity also harms Asian populations, with the risk for CKD onset being 1.13 times higher in men (95% confidence interval [CI]: 1.06– 1.20) and 1.02 times higher in women (95% CI: 0.97–1.07) compared with individuals of normal weight^2^. However, the body mass index (BMI) classification for defining obesity in Asians has been a persistent debate, with many experts arguing that the conventional threshold (BMI ≥30.0 kg/m^2^) is too high. This is because Asians typically have lower BMI but higher body fat percentage than Whites^3^. These racial differences in body composition are problematic when considering the relationship between BMI and CKD onset because higher body fat elevates the risk of CKD^4^. Thus, a BMI classification system with a lower cutoff might better reflect the relationship between obesity and CKD onset in Asian populations.

In 2000, the World Health Organization Western Pacific Region introduced the Asia-Pacific BMI classification (Asia-Pacific classification) for Asian adults^5^. This classification uses a lower BMI cutoff than the conventional World Health Organization BMI classification (conventional classification), which is applied across all races^6^. Follow-up studies utilizing the Asia-Pacific classification may help establish more accurate BMI targets for CKD prevention in Asians, but such studies remain limited.

Furthermore, diabetes complicates the relationship between obesity and CKD development^7^. A previous meta-analysis among Asians showed that obesity was associated with a 1.05-fold higher risk (95% CI: 1.01–1.10) of developing CKD in the diabetic population and a 1.06-fold higher risk (95% CI: 0.98–1.15) in the non-diabetic population compared with individuals of normal weight^2^. Several follow-up studies have statistically adjusted for prevalent diabetes or glycemic control status (e.g., fasting blood sugar levels) to minimize the confounding effect of diabetes^8–11^. However, diabetes-induced biases may remain because diabetes increases the risk of various factors that cause selection bias (e.g., coronary heart disease, ischemic stroke, and death)^12,13^. Therefore, longitudinal studies evaluating the relationship between obesity and the development of CKD in non-diabetic populations are important.

A previous cohort study of non-diabetic Japanese adults, the Ibaraki Prefecture Health Study (IPHS), revealed that BMI categories of ≥23.0 kg/m^2^ for men and ≥27.0 kg/m^2^ for women were associated with an elevated risk of CKD compared with the reference group^14,15^. However, this study independently categorized BMI into seven finer groups: <18.5, 18.5–20.9, 21.0–22.9 (reference), 23.0–24.9, 25.0–26.9, 27.0–29.9, and ≥30.0 kg/m^2^. Therefore, further follow-up studies using BMI categories standardized for Asians are needed to investigate the relationship between obesity and CKD onset in Asian populations^14^.

Based on the above, this longitudinal study investigated the relationship between obesity in the Asia-Pacific classification and the subsequent development of CKD in non-diabetic Japanese adults. By focusing on BMI thresholds specific to the Asian population, this study aims to address an important gap in CKD risk assessment.

## Materials and methods

### Data source

This study was a secondary data analysis using anonymized annual health checkup data from Zentsuji City (Kagawa Prefecture, Japan) from 6 April 1998 to 19 April 2023. As of 1 April 2023, the city had a population of 30,431 (49.7% male), with 38.1% of citizens aged ≥60 years^16^. The city database used in previous studies was used for data extraction^17–20^. Data extraction was conducted on 6 July 2023.

All citizens aged ≥40 years were eligible to receive a health checkup once every fiscal year, with participation being voluntary. Participants were free to join or leave the study at any time by choosing whether to undergo checkups. In fiscal years 1998 and 1999, younger citizens aged 34–39 years were also included on a trial basis to promote health awareness among younger populations. Over the years, approximately 30%–40% of the eligible population participated in the checkups^16^. No sampling procedures were implemented in this study.

During the checkups, the participants underwent anthropometric measurements, blood pressure tests, blood tests, and urine dipstick tests, and they completed a questionnaire regarding their lifestyle (e.g., alcohol intake, smoking status, medication use, and dietary and exercise habits)^21^. The checkups were conducted following the protocol established by the Ministry of Health, Labour and Welfare (MHLW), and all items were measured as specified in the protocol^21^. The checkups did not require participants to fast; therefore, they included both fasting and non-fasting individuals. Data on the participants’ last meal were not available.

### Study participants

This study evaluated the relationship between BMI and the development of CKD in non-diabetic Japanese adults. As such, it included all non-diabetic Japanese adults who underwent their first checkup between 1998 and 2023, were free of CKD at study entry, and had follow-up data to confirm the onset of CKD.

The following exclusion criteria were applied: non-Japanese individuals, individuals with missing exposure and/or outcome data, individuals with prevalent CKD and/or diabetes at study entry, individuals with missing hemoglobin A1c (HbA1c) values, individuals with prevalent diabetes at study entry, and individuals with only one observation (**Figure 1**). Participants who developed diabetes (HbA1c ≥ 6.5%) during follow-up were treated as right-censored^22^.

**Figure 1.**
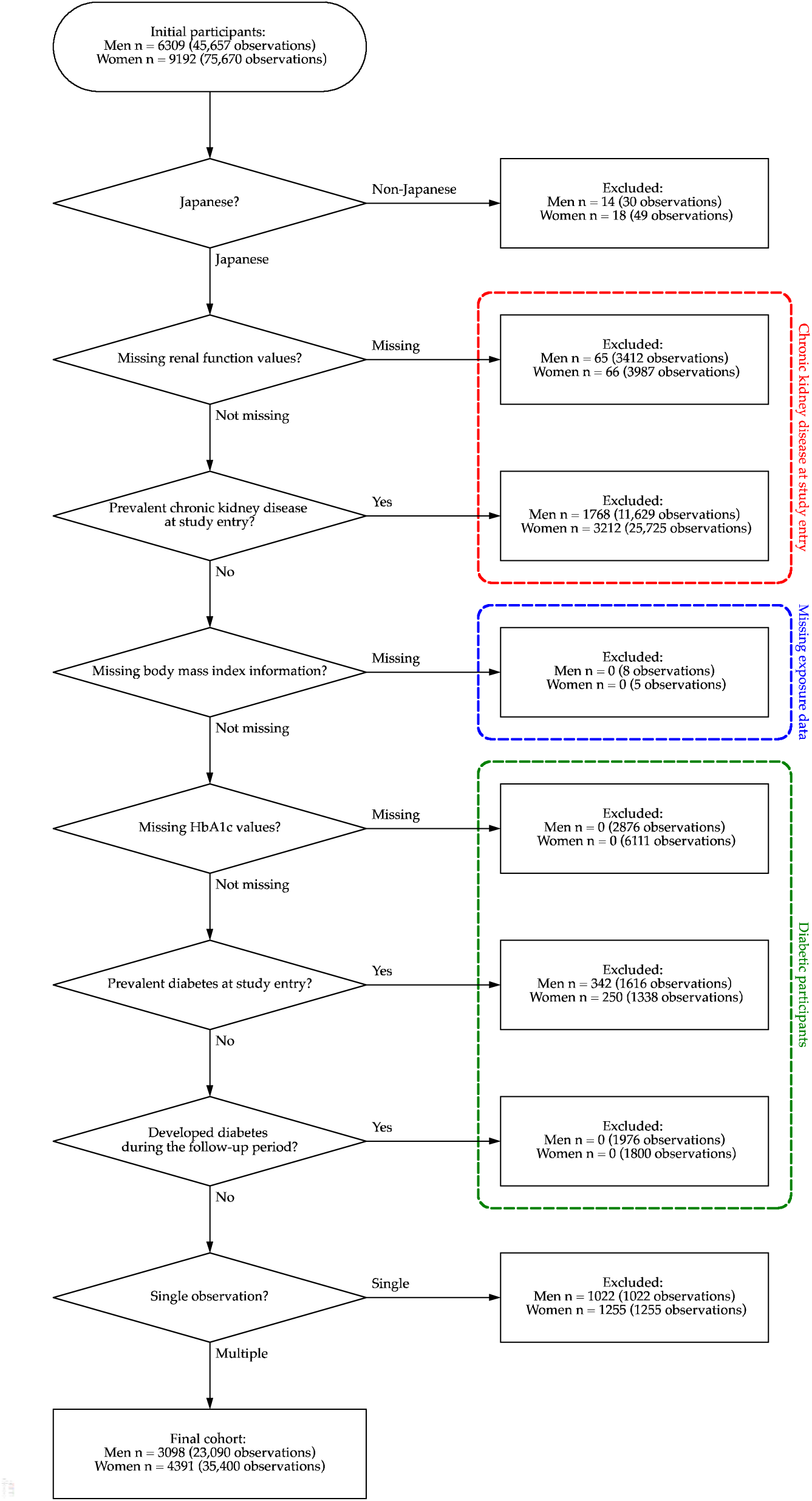
Participant flow chart of the study cohort.

### Variables

#### Exposure: BMI

The exposure variable was obesity, defined using BMI (calculated as weight in kilograms divided by height in meters squared) according to the Asia-Pacific classification: underweight (<18.5 kg/m^2^), normal weight (18.5– 22.9 kg/m^2^), overweight (23.0–24.9 kg/m^2^), obesity class I (25.0–29.9 kg/m^2^), and obesity class II (≥30.0 kg/m^2^)^5^. Obesity class II was categorized separately to assess its impact on the development of CKD in people with severe obesity and to avoid overestimating the effects of obesity class I by misattributing them to this category.

In this analysis, BMI classification was treated as a time-varying variable.

#### Outcome: CKD onset

The outcome variable was CKD onset, defined as an estimated glomerular filtration rate (eGFR) <60 mL/min/1.73 m^21^. Because this study exclusively included Japanese participants, eGFR was calculated using the three-variable revised Japanese equation: eGFR (mL/min/1.73m^2^) = 194 × serum creatinine (mg/dL)^-1.094^ × age (years)^-0.287^ (× 0.739, if female)^23^. Serum creatinine values were measured in mg/dL to two decimal places using the enzymatic method. Other information useful for determining CKD prevalence, such as definitive diagnoses by physicians, was unavailable and could not be included in this analysis^21^.

#### Other covariates

Sex was defined based on the participants’ physical characteristics at birth (male/female). Age (34–59 [reference]/60–69/70–100 years), self-reported alcohol intake (non- or seldom-drinker [reference]/drinker), self-reported smoking status (non- or ex-smoker [reference]/smoker), hypertension (no [reference]/yes), dyslipidemia (no [reference]/yes), HbA1c value (%), and eight residential districts (East [reference]/West/Central/South/Fudeoka/Tatsukawa/Yogita/Yoshiwara) were included as covariates for adjustment^1,2,24^.

Hypertension was defined as systolic blood pressure of ≥130 mmHg and/or diastolic blood pressure of ≥80 mmHg^25^. Dyslipidemia was defined as serum low-density lipoprotein cholesterol of ≥140 mg/dL, serum high-density lipoprotein cholesterol of <40 mg/dL, and/or serum triglycerides of ≥150 mg/dL^26^. HbA1c was reported using Japan Diabetes Society (JDS) units from 1998 to 2012 and National Glycohemoglobin Standardization Program (NGSP) units from 2013 to 2023^27,28^. HbA1c levels were standardized to NGSP values using the officially certified equation: HbA1c_NGSP_ (%) = 1.02 × HbA1c_JDS_ (%) + 0.25^29^. Neighborhood indicators (e.g., food quality and availability) have been reported to be associated with obesity, diabetes, and possibly CKD^24^. Therefore, to reduce the potential impact of the participants’ residential locations within the city, the district of residence was treated as a covariate.

This study had a substantial percentage of missing self-reported information (>50%) on medication use, dietary habits, and exercise habits, primarily due to changes in the questionnaire created by the MHLW^21^. A question regarding such information was added in 2012, but the question was either removed or modified in 2020 for older people aged ≥75 years^21^. To avoid age-related bias caused by this imbalance, these variables were excluded from the analysis.

### Statistical analysis

All analyses were conducted separately by sex. Participants’ characteristics were summarized by BMI class. Person-years at risk were calculated from the date of the first visit to the date of CKD or diabetes onset, or the end of the last visit during the follow-up period. Participant demographics during follow-up were expressed as numbers of patients with CKD onset, total person-years at risk, and incidence rate per 1,000 person-years.

We created Kaplan–Meier curves by the Asia-Pacific classification separately by sex. The proportional hazards assumption was violated as assessed by log–log plots and Schoenfeld residuals^30^. Therefore, the generalized gamma model was selected for the analysis based on the Akaike and Bayesian information criteria^30–33^. The time ratio and its 95% CI were used measures of the exposure effect on the outcome of interest. For example, a time ratio of 0.9 signified a 10% shorter survival time to CKD onset compared with the reference group.

Three adjusted models were constructed in addition to the crude model. Model 1 adjusted for age category. Model 2 further adjusted for self-reported alcohol intake and smoking status. Model 3 additionally adjusted for hypertension, dyslipidemia, HbA1c value, and residential district. To stabilize the models, a multiplicative term (BMI category × variable) was added to Models 1–3 if an interaction effect was observed between the BMI category and the following variables: age category, self-reported alcohol intake, self-reported smoking status, hypertension, dyslipidemia, or HbA1c values.

Because of the voluntary nature of the checkup, missing values were observed for several variables: self-reported alcohol intake (missing in 34.0% of men and 32.7% of women), self-reported smoking status (missing in 31.1% of men and 30.4% of women), hypertension (no missing data in men; missing in 0.02% of women), dyslipidemia (missing in 19.2% of men and 27.0% of women), and residential district (missing in 1.33% of men and 1.42% of women)^21^. Missing measurements were complemented using multiple imputation with chained equations, generating 40 imputations^34,35^. Binary variables and categorical variables were imputed using logistic regression and multinomial logistic regression, respectively. Only the imputed results were presented because missing measurements were assumed to be missing at random.

We restricted the presentation to the results for the main exposure variable to avoid the Table 2 fallacy^36^. A two-tailed p-value of <0.05 was considered statistically significant. All statistical analyses were performed using Stata/MP 16.1 (StataCorp, College Station, TX, USA). A participant flowchart was created in Python 3.11.10^37^. This study adhered to the Strengthening the Reporting of Observational Studies in Epidemiology (STROBE) reporting guidelines^38^.

### Sensitivity analyses

Three sensitivity analyses were conducted in this study. First, the conventional classification (underweight [<18.5 kg/m^2^], normal weight [18.5–24.9 kg/m^2^], overweight [25.0–29.9 kg/m^2^], obesity class I [30.0–34.9 kg/m^2^], obesity class II [35.0–39.9 kg/m^2^], and obesity class III [≥40.0 kg/m^2^]) was used as the exposure variable^6^. Because of the small number of participants with a BMI of ≥30.0 kg/m^2^, obesity classes I–III were combined into a single obesity category.

Second, to minimize the possibility of reverse causation, participants who developed CKD at the second observation were excluded. Third, to address the potential for temporary abnormal eGFR values, a stringent definition of CKD was applied. CKD was defined as two or more consecutive observations of eGFR <60 mL/min/1.73 m^2^. Given the use of annual checkup data, the duration of renal dysfunction in this stringent CKD definition was equal to or longer than the KDIGO definition of CKD, which requires a duration of ≥3 months^1^.

## Results

### Main analysis

Of 6,309 men and 9,192 women (mean age at study entry: 70.0 years for men, 68.3 years for women), 3,098 men and 4,391 women (mean age at study entry: 59.0 years for men, 62.1 years for women) were included in the final cohort (**Figure 1**). The mean follow-up time was slightly longer for women (8.25 years) than for men (7.41 years).

The participants’ characteristics by the Asia-Pacific BMI classification are presented in **Table 1** for men and **Table 2** for women. For both sexes, obesity class II had the shortest follow-up time, accounting for <3% of the total follow-up time. After the follow-up, 34.2% of men and 34.8% of women had developed CKD. Kaplan–Meier curves stratified by the Asia-Pacific classification, separately by sex, are provided in **Figure 2**. **Table 3** shows the results of the main analysis. Similar results were observed for both sexes, with obesity class I associated with an approximately 5% shorter survival time to CKD onset compared with normal weight.

**Figure 2.**
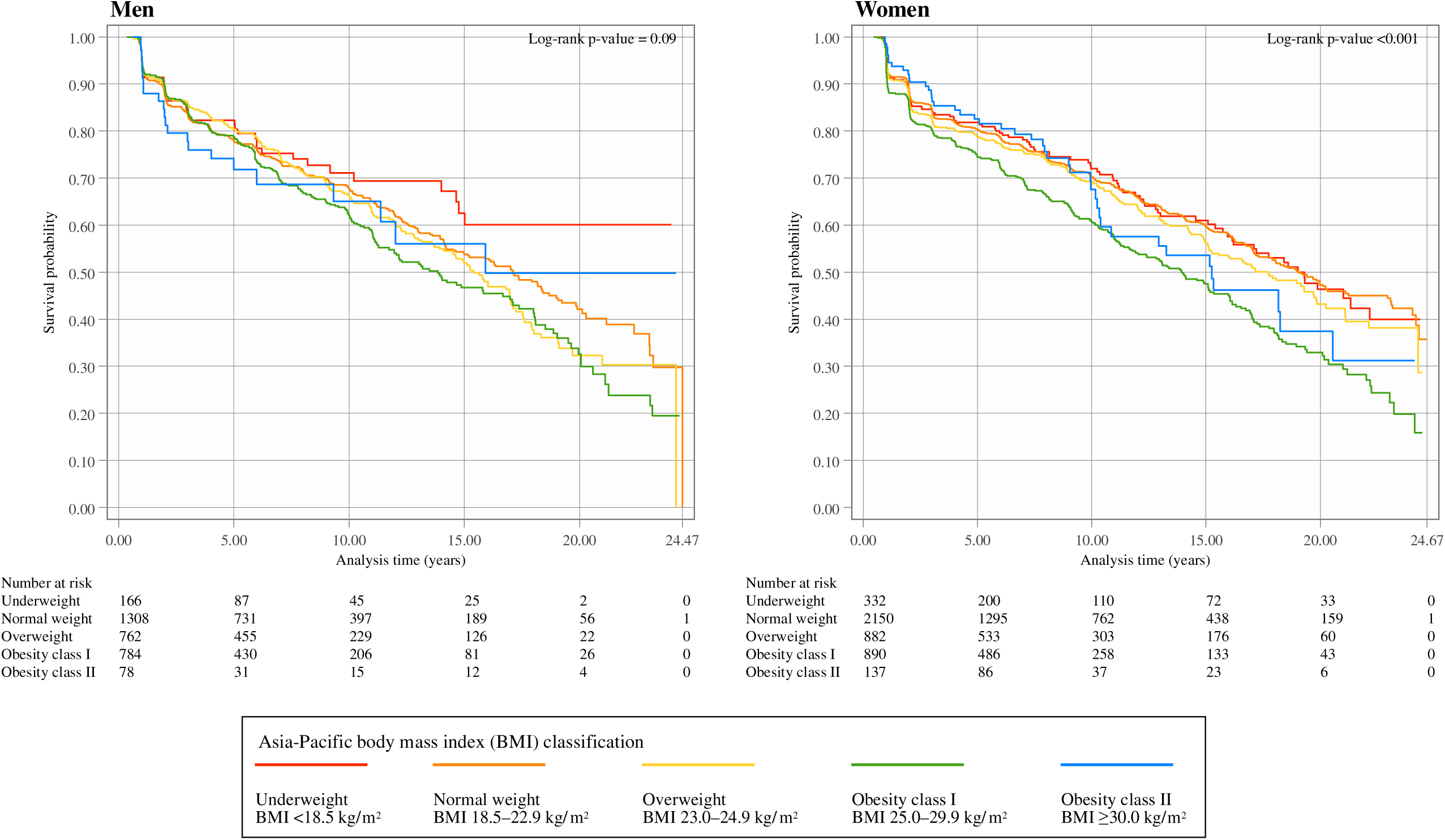
Kaplan–Meier survival estimates by Asia-Pacific body mass index classification for men and women.

**Table 1.**
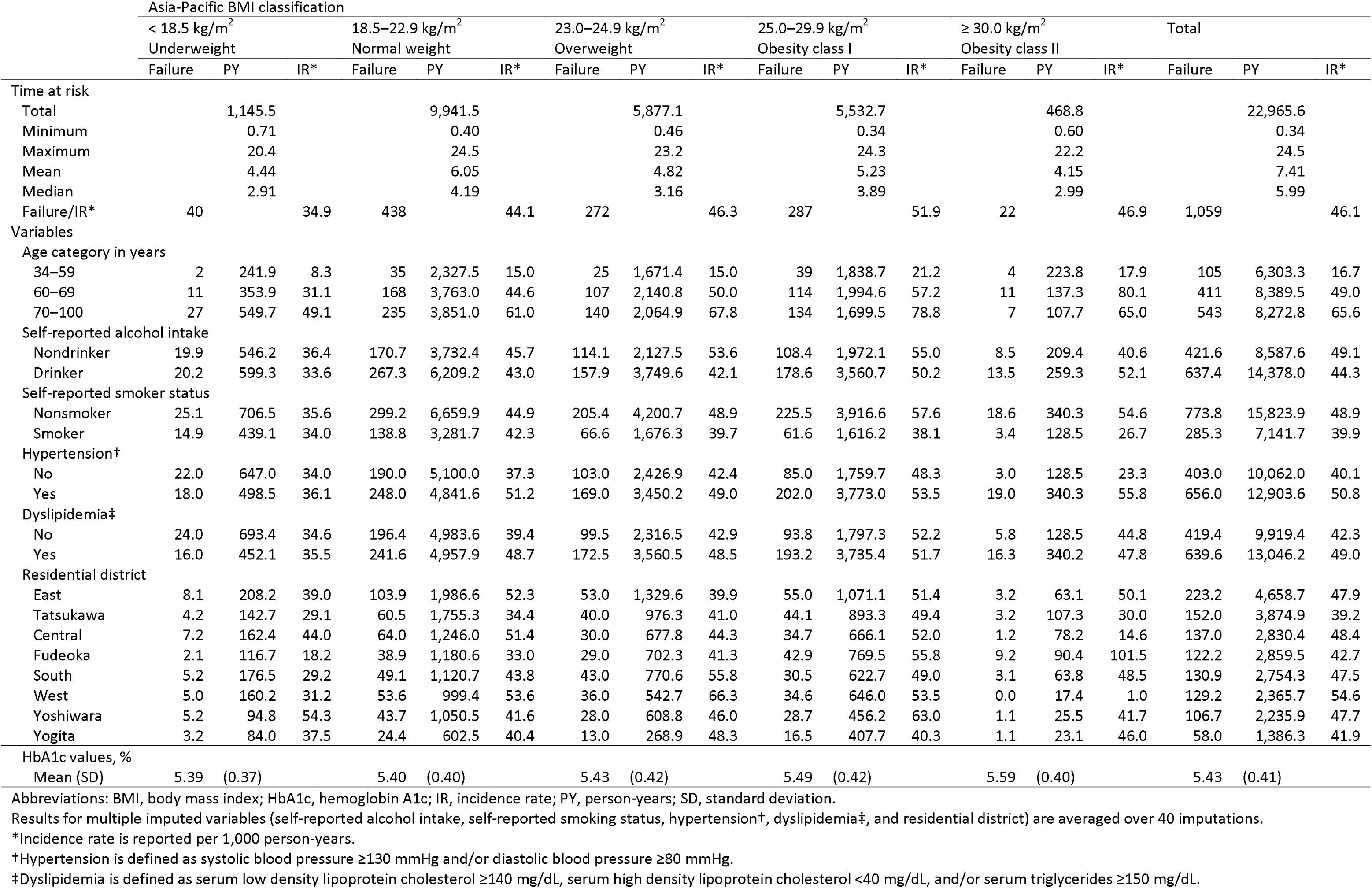
Descriptive statistics of all observations stratified by the Asia-Pacific BMI classification among 3,098 non-diabetic Japanese male citizens of Zentsuji City (1998–2023) Asia-Pacific BMI classification.

**Table 2.** Descriptive statistics of all observations stratified by the Asia-Pacific BMI classification among 4,391 non-diabetic Japanese female citizens of Zentsuji City (1998–2023)

|  | Asia-Pacific BMI classification |  |  |  |  |  |  |  |  |  |  |  |  |  |  |  |  |  |
| --- | --- | --- | --- | --- | --- | --- | --- | --- | --- | --- | --- | --- | --- | --- | --- | --- | --- | --- |
|  | < 18.5 kg/m<sup>2</sup> |  |  | 18.5–22.9 kg/m<sup>2</sup> |  |  | 23.0–24.9 kg/m<sup>2</sup> |  |  | 25.0–29.9 kg/m<sup>2</sup> |  |  | ≥ 30.0 kg/m<sup>2</sup> |  |  | Total |  |  |
|  | Underweight |  |  | Normal weight |  |  | Overweight |  |  | Obesity class I |  |  | Obesity class II |  |  |  |  |  |
|  | Failure | PY | IR* | Failure | PY | IR* | Failure | PY | IR* | Failure | PY | IR* | Failure | PY | IR* | Failure | PY | IR* |
| Time at risk |  |  |  |  |  |  |  |  |  |  |  |  |  |  |  |  |  |  |
| Total |  | 2,804.2 |  |  | 18,306.5 |  |  | 7,424.3 |  |  | 6,627.6 |  |  | 1,055.4 |  |  | 36,218.0 |  |
| Minimum |  | 0.63 |  |  | 0.48 |  |  | 0.48 |  |  | 0.53 |  |  | 0.80 |  |  | 0.48 |  |
| Maximum |  | 24.3 |  |  | 24.7 |  |  | 22.8 |  |  | 24.3 |  |  | 20.3 |  |  | 24.7 |  |
| Mean |  | 4.95 |  |  | 7.00 |  |  | 5.09 |  |  | 5.49 |  |  | 4.91 |  |  | 8.25 |  |
| Median |  | 3.05 |  |  | 5.05 |  |  | 3.63 |  |  | 3.87 |  |  | 3.10 |  |  | 6.83 |  |
| Failure/IR* | 106 |  | 37.8 | 697 |  | 38.1 | 307 |  | 41.4 | 371 |  | 56.0 | 45 |  | 42.6 | 1,526 |  | 42.1 |
| Variables |  |  |  |  |  |  |  |  |  |  |  |  |  |  |  |  |  |  |
| Age category in years |  |  |  |  |  |  |  |  |  |  |  |  |  |  |  |  |  |  |
| 34–59 | 23 | 1,137.7 | 20.2 | 187 | 7,825.2 | 23.9 | 70 | 2,392.4 | 29.3 | 73 | 1,980.3 | 36.9 | 9 | 436.8 | 20.6 | 362.0 | 13,772.4 | 26.3 |
| 60–69 | 23 | 761.2 | 30.2 | 199 | 5,690.4 | 35.0 | 93 | 2,816.1 | 33.0 | 117 | 2,455.0 | 47.7 | 12 | 300.7 | 39.9 | 444.0 | 12,023.4 | 36.9 |
| 70–100 | 60 | 905.2 | 66.3 | 311 | 4,790.9 | 64.9 | 144 | 2,215.9 | 65.0 | 181 | 2,192.3 | 82.6 | 24 | 317.9 | 75.5 | 720.0 | 10,422.2 | 69.1 |
| Self-reported alcohol intake |  |  |  |  |  |  |  |  |  |  |  |  |  |  |  |  |  |  |
| Nondrinker | 86.2 | 2,085.0 | 41.3 | 532.3 | 13,267.3 | 40.1 | 238.1 | 5,518.0 | 43.1 | 303.4 | 5,153.4 | 58.9 | 39.4 | 819.4 | 48.0 | 1,199.3 | 26,843.1 | 44.7 |
| Drinker | 19.9 | 719.1 | 27.6 | 164.7 | 5,039.3 | 32.7 | 68.9 | 1,906.4 | 36.2 | 67.6 | 1,474.2 | 45.8 | 5.7 | 236.0 | 23.9 | 326.7 | 9,375.0 | 34.8 |
| Self-reported smoker status |  |  |  |  |  |  |  |  |  |  |  |  |  |  |  |  |  |  |
| Nonsmoker | 102.6 | 2,568.8 | 39.9 | 668.3 | 17,266.9 | 38.7 | 297.0 | 7,090.5 | 41.9 | 362.5 | 6,334.9 | 57.2 | 43.8 | 991.1 | 44.2 | 1,474.3 | 34,252.2 | 43.0 |
| Smoker | 3.4 | 235.4 | 14.5 | 28.7 | 1,039.7 | 27.6 | 10.0 | 333.8 | 30.0 | 8.5 | 292.8 | 29.1 | 1.2 | 64.3 | 18.4 | 51.8 | 1,965.9 | 26.3 |
| Hypertension* |  |  |  |  |  |  |  |  |  |  |  |  |  |  |  |  |  |  |
| No | 53.0 | 1,950.8 | 27.2 | 330.0 | 10,753.6 | 30.7 | 110.0 | 3,377.2 | 32.6 | 113.0 | 2,314.1 | 48.8 | 11.0 | 243.3 | 45.2 | 617.0 | 18,639.0 | 33.1 |
| Yes | 53.0 | 853.3 | 62.1 | 367.0 | 7,553.0 | 48.6 | 197.0 | 4,047.2 | 48.7 | 258.0 | 4,313.5 | 59.8 | 34.0 | 812.0 | 41.9 | 909.0 | 17,579.0 | 51.7 |
| Dyslipidemia† |  |  |  |  |  |  |  |  |  |  |  |  |  |  |  |  |  |  |
| No | 64.6 | 1,776.0 | 36.4 | 330.1 | 9,347.6 | 35.3 | 119.3 | 3,067.8 | 38.9 | 126.4 | 2,521.8 | 50.1 | 16.5 | 404.0 | 40.8 | 656.9 | 17,117.1 | 38.4 |
| Yes | 41.5 | 1,028.1 | 40.4 | 366.9 | 8,959.0 | 41.0 | 187.7 | 4,356.6 | 43.1 | 244.6 | 4,105.8 | 59.6 | 28.5 | 651.4 | 43.8 | 869.1 | 19,100.9 | 45.5 |
| Residential district |  |  |  |  |  |  |  |  |  |  |  |  |  |  |  |  |  |  |
| East | 22.0 | 508.0 | 43.3 | 132.8 | 3,621.6 | 36.7 | 57.8 | 1,429.9 | 40.4 | 76.7 | 1,447.1 | 53.0 | 8.0 | 215.2 | 37.2 | 297.3 | 7,221.8 | 41.2 |
| Tatsukawa | 15.0 | 557.7 | 26.9 | 101.0 | 3,329.9 | 30.3 | 57.0 | 1,223.3 | 46.6 | 52.6 | 1,102.7 | 47.7 | 7.0 | 170.3 | 41.1 | 232.6 | 6,383.9 | 36.4 |
| Central | 21.0 | 452.7 | 46.4 | 106.5 | 2,508.4 | 42.5 | 43.0 | 950.6 | 45.2 | 51.3 | 874.0 | 58.7 | 11.0 | 157.2 | 70.0 | 232.8 | 4,943.0 | 47.1 |
| Fudeoka | 10.0 | 278.3 | 35.9 | 93.4 | 2,344.1 | 39.8 | 31.8 | 954.4 | 33.3 | 49.5 | 858.1 | 57.7 | 3.0 | 146.8 | 20.4 | 187.7 | 4,581.7 | 41.0 |
| South | 11.0 | 363.7 | 30.3 | 70.1 | 1,917.1 | 36.6 | 31.8 | 808.7 | 39.3 | 39.5 | 702.6 | 56.3 | 4.0 | 109.7 | 36.5 | 156.4 | 3,901.8 | 40.1 |
| West | 6.0 | 292.9 | 20.5 | 69.0 | 1,903.6 | 36.3 | 38.7 | 791.4 | 48.8 | 43.4 | 586.3 | 74.1 | 5.0 | 111.0 | 45.1 | 162.1 | 3,685.2 | 44.0 |
| Yoshiwara | 10.0 | 192.0 | 52.1 | 80.8 | 1,773.7 | 45.5 | 28.6 | 819.1 | 34.9 | 35.6 | 591.0 | 60.2 | 4.0 | 96.0 | 41.7 | 159.0 | 3,471.8 | 45.8 |
| Yogita | 11.0 | 158.7 | 69.3 | 43.5 | 908.1 | 47.8 | 18.4 | 447.0 | 41.1 | 22.4 | 465.8 | 48.0 | 3.0 | 49.3 | 61.0 | 98.2 | 2,029.0 | 48.4 |
| HbA1c values, % |  |  |  |  |  |  |  |  |  |  |  |  |  |  |  |  |  |  |
| Mean (SD) | 5.45 | (0.34) |  | 5.45 | (0.38) |  | 5.50 | (0.39) |  | 5.53 | (0.39) |  | 5.54 | (0.37) |  | 5.48 | (0.38) |  |

Abbreviations: BMI, body mass index; HbA1c, hemoglobin A1c; IR, incidence rate; PY, person-years; SD, standard deviation.
Results for multiple imputed variables (self-reported alcohol intake, self-reported smoking status, hypertension\*, dyslipidemia†, and residential district) are averaged over 40 imputations.
\*Incidence rate is reported per 1,000 person-years.
†Hypertension is defined as systolic blood pressure ≥130 mmHg and/or diastolic blood pressure ≥80 mmHg.
‡Dyslipidemia is defined as serum low density lipoprotein cholesterol ≥140 mg/dL, serum high density lipoprotein cholesterol <40 mg/dL, and/or serum triglycerides ≥150 mg/dL.

**Table 3.** New onset of chronic kidney disease defined by the Asia-Pacific BMI classification in non-diabetic Japanese citizens of Zentsuji City (1998–2023)

| Asia-Pacific BMI classification |  | PY | Failure | IR‡ | Crude<br>TR (95% CI) | Model 1<br>aTR (95% CI) | Model 2<br>aTR (95% CI) | Model 3<br>aTR (95% CI) |
| --- | --- | --- | --- | --- | --- | --- | --- | --- |
| **Men (n=3,098)** |  |  |  |  |  |  |  |  |
| <18.5 kg/m<sup>2</sup> | Underweight | 1,145.5 | 40 | 34.9 | 1.08 (1.00–1.16) | 1.08 (0.99–1.17) | 1.08 (0.99–1.17) | 1.08 (1.00–1.17) |
| 18.5–22.9 kg/m<sup>2</sup> (reference) | Normal weight | 9,941.5 | 438 | 44.1 | 1.00 | 1.00 | 1.00 | 1.00 |
| 23.0–24.9 kg/m<sup>2</sup> | Overweight | 5,877.1 | 272 | 46.3 | 0.97 (0.94–1.01) | 0.97 (0.94–1.00) | 0.97 (0.93–1.00) | 0.97 (0.94–1.01) |
| 25.0–29.9 kg/m<sup>2</sup> | Obesity class I | 5,532.7 | 287 | 51.9 | 0.94 (0.90–0.97) | 0.93 (0.90–0.97) | 0.93 (0.90–0.97) | 0.94 (0.90–0.98) |
| ≥30.0 kg/m<sup>2</sup> | Obesity class II | 468.8 | 22 | 46.9 | 0.92 (0.82–1.02) | 0.92 (0.82–1.02) | 0.92 (0.82–1.02) | 0.92 (0.82–1.02) |
| Asia-Pacific BMI classification |  | PY | Failure | IR‡ | Crude<br>TR (95% CI) | Model 1<br>aTR (95% CI) | Model 2<br>aTR (95% CI) | Model 3<br>aTR (95% CI) |
| **Women (n=4,391)** |  |  |  |  |  |  |  |  |
| <18.5 kg/m<sup>2</sup> | Underweight | 2,804.2 | 106 | 37.8 | 1.02 (0.97–1.06) | 1.02 (0.98–1.06) | 1.02 (0.97–1.06) | 1.01 (0.96–1.05) |
| 18.5–22.9 kg/m<sup>2</sup> (reference) | Normal weight | 18,306.5 | 697 | 38.1 | 1.00 | 1.00 | 1.00 | 1.00 |
| 23.0–24.9 kg/m<sup>2</sup> | Overweight | 7,424.3 | 307 | 41.4 | 0.99 (0.96–1.02) | 0.99 (0.96–1.02) | 0.99 (0.96–1.02) | 1.00 (0.97–1.03) |
| 25.0–29.9 kg/m<sup>2</sup> | Obesity class I | 6,627.6 | 371 | 56.0 | 0.94 (0.92–0.97) | 0.95 (0.92–0.97) | 0.95 (0.92–0.97) | 0.95 (0.93–0.98) |
| ≥30.0 kg/m<sup>2</sup> | Obesity class II | 1,055.4 | 45 | 42.6 | 0.97 (0.91–1.03) | 0.97 (0.91–1.03) | 0.97 (0.91–1.03) | 0.98 (0.92–1.05) |

Abbreviations: aTR, adjusted time ratio; BMI, body mass index; CI, confidence interval; HbA1c, hemoglobin A1c; IR, incidence rate; PY, person-years; TR, time ratio.
\*Hypertension is defined as systolic blood pressure ≥130 mmHg and/or diastolic blood pressure ≥80 mmHg.
†Dyslipidemia is defined as serum low-density lipoprotein cholesterol ≥140 mg/dL, serum high-density lipoprotein cholesterol <40 mg/dL, and/or serum triglycerides ≥150 mg/dL.
‡Incidence rate is reported per 1,000 person-years.
Multiple imputed variables: hypertension\*, dyslipidemia†, self-reported alcohol intake, self-reported smoking status, and residential district.
Model 1: Adjusted for age category (34–59[reference]/60–69/70–100).
Model 2: Adjusted for the variable of Model 1, self-reported alcohol intake (non- or seldom-drinker [reference]/drinker), and self-reported smoking status (non- or ex-smoker [reference]/smoker).
Model 3: Adjusted for all variables of Model 2, hypertension\* (no[reference]/yes), dyslipidemia† (no[reference]/yes), HbA1c values, and residential district (East[reference]/Tatsukawa/Central/Fudeoka/South/West/Yoshiwara/Yogita).

For men, obesity class II had the shortest survival time to CKD onset, with a point estimate of 8%, but the 95% CIs included 1 in all models. For women in obesity class II, the results were nearly null.

For reference, using the combined BMI categories of the top two categories (obesity class I and II) as a single obesity category yielded results similar to those of the main analysis: the adjusted time ratios (95% CIs) of overweight and obesity were 0.97 (0.94–1.01) and 0.94 (0.90–0.97) for men and 1.00 (0.97–1.03) and 0.95 (0.93–0.98) for women.

### Sensitivity analyses

#### First sensitivity analysis: The conventional classification

Using the conventional classification, more participants (men: 68.9%, women: 71.0%) were classified as normal weight than in the Asia-Pacific classification (men: 43.3%, women: 50.6%) (**Tables 1 and 2**)^6^. The estimation results were similar for a BMI of ≥30.0 kg/m^2^ (“obesity class II” in the Asia-Pacific classification and “obesity” in the conventional classification), showing shorter survival to CKD development in higher BMI categories (**Tables 3 and S1**)^5,6^.

#### Second sensitivity analysis: Reverse causation

The second sensitivity analysis aimed to minimize reverse causation by excluding participants who developed CKD at the second observation, resulting in 2,723 men and 3,832 women remaining in the cohort. The mean age at study entry was higher for men (61.4 years) and lower for women (58.3 years) compared with the main analysis. The mean follow-up period was 8.14 years for men and 9.14 years for women, slightly longer than in the main analysis (**Tables 1 and 2**). The distribution of follow-up time by BMI category was similar to that of the main analysis.

After the follow-up period, 25.1% of men and 25.2% of women had developed CKD, lower than in the main analysis. Most estimation results were consistent with those of the main analysis; however, the point estimates of survival for obesity class II were longer in men and shorter in women compared with the main analysis (**Tables 3 and S2**).

#### Third sensitivity analysis: Stringent definition of CKD

To minimize variability in eGFR measurements, a stringent definition of CKD requiring two consecutive observations of eGFR <60 mL/min/1.73 m^2^ was applied (**Table S3**). Compared with the main analysis (**Tables 1 and 2**), more participants (men: 3,380; women: 4,963) remained in the final cohort, the mean follow-up time was longer (men: 8.13 years; women: 9.29 years), the mean age at study entry was higher for men (62.6 years) and lower for women (59.6 years), and fewer participants developed CKD (men: 20.7%; women: 21.1%).

For men, point estimates for higher BMI categories were similar to those in the main analysis (**Table 3**), including obesity class II, which had the shortest follow-up period. For women, convergence errors were observed in Model 3. The results of Model 2 for women were similar to those of Model 2 in the main analysis.

## Discussion

### Main analysis

In this population-based longitudinal study (1998–2023) of non-diabetic Japanese adults, obesity class I in the Asia-Pacific classification was associated with earlier CKD onset. Although the 95% CIs for overweight and obesity class II included 1 in all models, the point estimates suggested a dose–response relationship, particularly in men (**Table 3**).

These findings partly align with those of the previous non-diabetic Japanese cohort study from the IPHS^14,15^. Similar to our study, the IPHS demonstrated a dose–response relationship between BMI and CKD onset: CKD risk was higher in BMI categories 23.0–24.9, 25.0–26.9, 27.0–29.9, and ≥30.0 kg/m^2^ in men and 27.0–29.9 and ≥30.0 kg/m^2^ in women compared with the BMI category 21.0–22.9 kg/m^2^. However, our study showed a dose–response relationship by point estimates in men but not in women.

Three factors may explain this discrepancy. First, the sample size in this study (n = 7,489) was smaller than that of the IPHS (n = 89,760)^14^, potentially resulting in insufficient power to detect a true effect in the small proportion of participants with obesity class II (<3% of the total population). Second, our participants were older (range: 34–100 years) than those in the IPHS (range: 40–79 years). To mitigate bias, adjustments for age were made to reduce the likelihood of over- or underestimation caused by older adults’ tendency to have a lower BMI^39^. Nonetheless, the generalizability of our findings to populations with different age distributions remains limited. Third, the observational period in this study (1998–2023) was longer than that of the IPHS (1993–2006). A longer follow-up period may decrease participants’ susceptibility to outcomes over time^40^. Specifically, the extended observational period in our study likely introduced more selection bias compared with the IPHS, contributing to an underestimation of results^40^.

In this study, men had a shorter survival time to CKD onset than women with the same BMI (**Tables 1 and 2**). This trend was consistent with the findings from the IPHS and can be partly attributed to biological and physiological differences between sexes^14^. Higher muscle mass in men than in women may result in systematic errors in estimation of the GFR because higher muscle mass can be misclassified as a lower eGFR^41,42^. Consequently, the results are likely to be overestimated in men. However, most participants in this study were older adults (i.e., those aged ≥70 years), who tend to have a lower BMI and muscle mass, leading to underestimation (**Tables 1 and 2**)^39,41^. Therefore, the sex-related overestimation is likely attenuated. Furthermore, as noted above, this study had large random errors in obesity class II, which accounted for the shortest follow-up time (<3% of the total follow-up period) (**Tables 1 and 2**). As a result, unmeasured factors may have had a greater impact on the results of this study, especially in obesity class II.

### Sensitivity analyses

#### First sensitivity analysis: Conventional classification

**Table S1** presents the results using the conventional BMI classification as the exposure variable^6^. Obesity had the lowest point estimates for men and an almost null value for women, with all 95% CIs crossing 1. Compared with the findings of the main analysis, overweight (which corresponds to obesity class I in the Asia-Pacific classification) was associated with shorter survival to CKD onset. These results suggest that the conventional BMI cutoff of ≥30.0 kg/m^2^ for obesity may be too high to act as a risk factor for CKD in non-diabetic Japanese adult populations.

#### Second sensitivity analysis: Reverse causation

**Table S2** shows the results after minimizing the possibility of reverse causation by excluding participants who developed CKD at the second observation. For all BMI categories below 30.0 kg/m^2^, the results followed a trend similar to that observed in the main analysis (**Table 3**), suggesting no clear evidence of reverse causation between these BMI categories and CKD onset.

However, a weak inverse association was observed in the BMI ≥30.0 kg/m^2^ category. As noted earlier, evaluating the relationship between BMI ≥30.0 kg/m^2^ and CKD onset was hindered by reduced statistical power. Consequently, a type II error may have occurred in this category, suggesting the possibility of a dose– response relationship between BMI and CKD development in this analysis.

#### Third sensitivity analysis: Stringent definition of CKD

Using the stringent CKD definition, where two consecutive observations of eGFR <60 mL/min/1.73 m^2^ were required to define CKD, the results (**Table S3**) showed trends similar to the main analysis in Models 1 and 2 (**Table 3**). Compared with the main analysis (men: 1,059; women: 1,526), fewer participants developed CKD (men: 700; women: 1,049), which caused a convergence error in Model 3.

### Strengths and limitations

A strength of this study was its use of the Asia-Pacific BMI classification to evaluate the impact of elevated BMI on the development of CKD in non-diabetic monoracial Asian men and women, using longitudinal data with a relatively long follow-up period (1998–2023).

However, several limitations must be noted. First, because East Asians tend to have a lower BMI than Westerners, the number of participants in the highest BMI category (≥30.0 kg/m^2^) was low in this study, and the relationship between BMI and subsequent development of CKD could not be fully assessed^3,43^.

Second, a high BMI does not necessarily equate to obesity. The BMI is calculated based on weight and height and does not account for body composition, such as body fat percentage. When obesity is defined as body fat percentages of ≥25% for men and ≥35% for women in an Asian population, detecting obesity using a BMI of ≥30.0 kg/m^2^ has high specificity (men: 97.3%, women: 95.0%) but low sensitivity (men: 6.7%, women: 13.4%)^44^. Therefore, a BMI cutoff of ≥30.0 kg/m^2^ misses most participants with high body fat, resulting in misclassification and underestimation in the highest BMI category^3,43^.

Third, the exact date of CKD onset could not be determined—a major and unresolved limitation of cohort studies that are based on annual checkup data and in which participants can decide when to be screened. These factors lead to interval censoring, which requires highly complex statistical models to address^45^.

Fourth, measurement errors in renal function may have occurred. Because this study was a secondary data analysis, inulin clearance values (the gold standard for assessing GFR) were unavailable for evaluation^1^. Instead, we used serum creatinine to estimate GFR using the three-variable revised Japanese equation, which is more accurate than the Modification of Diet in Renal Disease equations^23^. Furthermore, other useful indicators for detecting CKD (e.g., cystatin C) were also unavailable in this study^1^. The inability to combine such variables to define CKD may have resulted in an overestimation of the association ^1,46^.

Fifth, the results of this study cannot be generalized because the data were not obtained through random sampling from the general population. Furthermore, because only 30%–40% of the target population participated in the checkups, the generalizability of the results is further limited^16^. Furthermore, our study participants were likely healthier than the general population because people who voluntarily attend checkups have been reported to be more health-conscious and healthier^47^. This may introduce selection bias and underestimate the findings.

Sixth, competing risks that may have modified the occurrence of events (e.g., death, hospitalization) could not be assessed because the necessary data were unavailable. Finally, indicators of central and visceral fat (e.g., waist circumference, waist-to-hip ratio) were not available. Among 454 adult Koreans, elevations in these factors were more strongly associated with CKD than BMI regardless of the diabetes status^48^. In addition, one review showed that the waist-to-height ratio is a better indicator of obesity and is associated with CKD^49^. Future studies should incorporate these variables alongside BMI to create an obesity index that better predicts CKD risk by race for both sexes without diabetes.

## Conclusion

Our study of 7,489 non-diabetic Japanese adults, using the Asia-Pacific BMI classification, showed that both sexes in obesity class I had approximately 5% shorter survival to subsequent CKD onset compared with those of normal weight. Obesity class II had the shortest survival to CKD onset by point estimates in men and an almost null value in women, with all 95% CIs crossing 1. These results align with previous findings of a dose– response relationship between BMI and CKD development, particularly in men. Our study provides additional evidence suggesting that the BMI cutoff values for obesity, as defined by the conventional BMI classification, may be too high when evaluating the association between obesity and future CKD development in non-diabetic Japanese adults. Our findings may serve as a useful indicator when identifying the target population for future CKD prevention efforts.

## Ethics

All data were anonymized before receipt. The Ethics Committee of Okayama University Graduate School of Medicine, Dentistry and Pharmaceutical Sciences and Okayama University Hospital approved this study (No. K1708-040). The Ethics Committee waived the need for informed consent. Study procedures were performed in accordance with the Declaration of Helsinki and Japanese Ethical Guidelines for Medical and Biological Research Involving Human Subjects.

## Data Availability

All data generated or analyzed during this study are included in this published article and its supplementary information files.

## Acknowledgments

We are grateful to all participants of this study, Ayaka Nakatsu, Masako Matsumoto, Mayumi Kitadani, and all local government officers of Zentsuji City for their support and contribution. We thank Anahid Pinchis, BSc, MBA, Angela Morben, DVM, and Ellen Knapp, PhD, from Edanz (https://jp.edanz.com/ac) for editing a draft of this manuscript.

