## Supplementary Tables for "The Asia-Pacific body mass index classification and new-onset chronic kidney disease in non-diabetic Japanese adults: A community-based longitudinal study from 1998 to 2023"

**Table S1**. New onset of chronic kidney disease defined by the conventional WHO BMI classification in non-diabetic Japanese citizens of Zentsuji City (1998–2023)

| Conventional WHO BMI classification | | Person-years | Failure | IR‡ | Crude TR (95% CI) | Model 1 aTR (95% CI) | Model 2 aTR (95% CI) | Model 3 aTR (95% CI) |
|---|---|---|---|---|---|---|---|---|
| **Men (n=3,098)** | | | | | | | | |
| <18.5 kg/m$^2$ | Underweight | 1,145.5 | 40 | 34.9 | 1.09 (1.01–1.18) | 1.09 (1.00–1.18) | 1.09 (1.01–1.19) | 1.09 (1.01–1.19) |
| 18.5–24.9 kg/m$^2$ (reference) | Normal weight | 15,818.6 | 710 | 44.9 | 1.00 | 1.00 | 1.00 | 1.00 |
| 25.0–29.9 kg/m$^2$ | Overweight | 5,532.7 | 287 | 51.9 | 0.94 (0.91–0.98) | 0.94 (0.91–0.97) | 0.94 (0.91–0.97) | 0.95 (0.92–0.98) |
| ≥30.0 kg/m$^2$ | Obesity | 468.8 | 22 | 46.9 | 0.92 (0.83–1.03) | 0.93 (0.83–1.03) | 0.93 (0.83–1.03) | 0.93 (0.83–1.03) |
| Conventional WHO BMI classification | | Person-years | Failure | IR‡ | Crude TR (95% CI) | Model 1 aTR (95% CI) | Model 2 aTR (95% CI) | Model 3 aTR (95% CI) |
| **Women (n=4,391)** | | | | | | | | |
| <18.5 kg/m$^2$ | Underweight | 2,804.2 | 106 | 37.8 | 1.02 (0.98–1.07) | 1.02 (0.98–1.06) | 1.02 (0.98–1.06) | 1.01 (0.96–1.05) |
| 18.5–24.9 kg/m$^2$ (reference) | Normal weight | 25,730.9 | 1,004 | 39.0 | 1.00 | 1.00 | 1.00 | 1.00 |
| 25.0–29.9 kg/m$^2$ | Overweight | 6,627.6 | 371 | 56.0 | 0.95 (0.93–0.97) | 0.95 (0.93–0.97) | 0.95 (0.93–0.97) | 0.95 (0.93–0.98) |
| ≥30.0 kg/m$^2$ | Obesity | 1,055.4 | 45 | 42.6 | 0.97 (0.91–1.03) | 0.97 (0.91–1.03) | 0.97 (0.91–1.03) | 0.98 (0.92–1.05) |

Abbreviations: aTR, adjusted time ratio; BMI, body mass index; CI, confidence interval; HbA1c, hemoglobin A1c; IR, incidence rate; TR, time ratio, WHO, World Health Organization.

*Hypertension is defined as systolic blood pressure ≥130 mmHg and/or diastolic blood pressure ≥80 mmHg.

†Dyslipidemia is defined as serum low-density lipoprotein cholesterol ≥140 mg/dL, serum high-density lipoprotein cholesterol <40 mg/dL, and/or serum triglycerides ≥150 mg/dL.

‡Incidence rate is reported per 1,000 person-years.

Multiple imputed variables: hypertension*, dyslipidemia†, self-reported alcohol intake, self-reported smoking status, and residential district.

Model 1: Adjusted for age category (34–59[reference]/60–69/70–100).

Model 2: Adjusted for the variable of Model 1, self-reported alcohol intake (non- or seldom-drinker [reference]/drinker), and self-reported smoking status (non- or ex-smoker [reference]/smoker).

Model 3: Adjusted for all variables of Model 2, hypertension* (no[reference]/yes), dyslipidemia† (no[reference]/yes), HbA1c values, and residential district (East[reference]/Tatsukawa/Central/Fudeoka/South/West/Yoshiwara/Yogita).

**Table S2**. New onset of CKD defined by the Asia-Pacific BMI classification in non-diabetic Japanese citizens of Zentsuji City, with exclusion of those who had developed CKD at the second observation (1998–2023)

| | | | | | Crude | Model 1 | Model 2 | Model 3 |
|---|---|---|---|---|---|---|---|---|
| Asia-Pacific BMI classification | | Person-years | Failure | IR‡ | TR (95% CI) | aTR (95% CI) | aTR (95% CI) | aTR (95% CI) |
| **Men (n=2,723)** | | | | | | | | |
| <18.5 kg/m$^2$ | Underweight | 1,128.6 | 25 | 22.2 | 1.08 (1.00–1.17) | 1.08 (1.00–1.17) | 1.08 (1.00–1.17) | 1.08 (1.00–1.17) |
| 18.5–22.9 kg/m$^2$ (reference) | Normal weight | 9,646.1 | 284 | 29.4 | 1.00 | 1.00 | 1.00 | 1.00 |
| 23.0–24.9 kg/m$^2$ | Overweight | 5,648.0 | 177 | 31.3 | 0.97 (0.94–1.01) | 0.97 (0.94–1.01) | 0.97 (0.94–1.01) | 0.98 (0.94–1.01) |
| 25.0–29.9 kg/m$^2$ | Obesity class I | 5,305.7 | 188 | 35.4 | 0.94 (0.91–0.97) | 0.94 (0.90–0.97) | 0.94 (0.90–0.97) | 0.94 (0.91–0.98) |
| ≥30.0 kg/m$^2$ | Obesity class II | 449.9 | 10 | 22.2 | 0.98 (0.88–1.10) | 0.98 (0.88–1.10) | 0.98 (0.88–1.10) | 0.98 (0.88–1.10) |
| | | | | | Crude | Model 1¶ | Model 2¶ | Model 3§ |
| Asia-Pacific BMI classification | | Person-years | Failure | IR‡ | TR (95% CI) | aTR (95% CI) | aTR (95% CI) | aTR (95% CI) |
| **Women (n=3,832)** | | | | | | | | |
| <18.5 kg/m$^2$ | Underweight | 2,718.9 | 65 | 23.9 | 1.03 (0.98–1.07) | 1.06 (0.88–1.28) | 1.06 (0.88–1.27) | 1.07 (1.00–1.16) |
| 18.5–22.9 kg/m$^2$ (reference) | Normal weight | 17,753.1 | 449 | 25.3 | 1.00 | 1.00 | 1.00 | 1.00 |
| 23.0–24.9 kg/m$^2$ | Overweight | 7,156.6 | 188 | 26.3 | 1.00 (0.97–1.03) | 0.94 (0.85–1.05) | 0.94 (0.85–1.05) | 0.99 (0.94–1.04) |
| 25.0–29.9 kg/m$^2$ | Obesity class I | 6,356.4 | 232 | 36.5 | 0.96 (0.93–0.98) | 0.88 (0.80–0.96) | 0.88 (0.80–0.96) | 0.92 (0.88–0.96) |
| ≥30.0 kg/m$^2$ | Obesity class II | 1,027.6 | 33 | 32.1 | 0.96 (0.90–1.02) | 0.92 (0.74–1.14) | 0.92 (0.74–1.14) | 0.92 (0.83–1.02) |

Abbreviations: aTR, adjusted time ratio; BMI, body mass index; CI, confidence interval; CKD, chronic kidney disease; HbA1c, hemoglobin A1c; IR, incidence rate; TR, time ratio.

*Hypertension is defined as systolic blood pressure ≥130 mmHg and/or diastolic blood pressure ≥80 mmHg.

†Dyslipidemia is defined as serum low-density lipoprotein cholesterol ≥140 mg/dL, serum high-density lipoprotein cholesterol <40 mg/dL, and/or serum triglycerides ≥150 mg/dL.

‡Incidence rate is reported per 1,000 person-years.

Multiple imputed variables: hypertension*, dyslipidemia†, self-reported alcohol intake, self-reported smoking status, and residential district.

Model 1: Adjusted for age category (34–59[reference]/60–69/70–100).

Model 2: Adjusted for the variable of Model 1, self-reported alcohol intake (non- or seldom-drinker [reference]/drinker), and self-reported smoking status (non- or ex-smoker [reference]/smoker).

Model 3: Adjusted for all variables of Model 2, hypertension* (no[reference]/yes), dyslipidemia† (no[reference]/yes), HbA1c values, and residential district (East[reference]/Tatsukawa/Central/Fudeoka/South/West/Yoshiwara/Yogita).

¶A multiplicative term (BMI classification × age category) was added.

§A multiplicative term (BMI classification × hypertension*) was added.

**Table S3**. New onset of CKD defined by the Asia-Pacific BMI classification in non-diabetic Japanese citizens of Zentsuji City using a stringent CKD definition (at least two consecutive observations of an eGFR <60 mL/min/1.73 $m^2$ is considered CKD) (1998–2023)

| Asia-Pacific BMI classification | | Person-years | Failure | IR‡ | Crude | Model 1 | Model 2 | Model 3 |
|---|---|---|---|---|---|---|---|---|
| | | | | | TR (95% CI) | aTR (95% CI) | aTR (95% CI) | aTR (95% CI) |
| **Men (n=3,380)** | | | | | | | | |
| <18.5 kg/$m^2$ | Underweight | 1,364.1 | 29 | 21.3 | 1.04 (0.96–1.13) | 1.03 (0.96–1.11) | 1.03 (0.96–1.11) | 1.03 (0.96–1.11) |
| 18.5–22.9 kg/$m^2$ (reference) | Normal weight | 12,009.1 | 274 | 22.8 | 1.00 | 1.00 | 1.00 | 1.00 |
| 23.0–24.9 kg/$m^2$ | Overweight | 7,008.4 | 192 | 27.4 | 0.95 (0.92–0.99) | 0.95 (0.92–0.99) | 0.95 (0.92–0.99) | 0.96 (0.93–0.99) |
| 25.0–29.9 kg/$m^2$ | Obesity class I | 6,560.5 | 189 | 28.8 | 0.93 (0.89–0.96) | 0.93 (0.90–0.97) | 0.94 (0.90–0.97) | 0.94 (0.91–0.98) |
| ≥30.0 kg/$m^2$ | Obesity class II | 538.3 | 16 | 29.7 | 0.88 (0.79–0.98) | 0.89 (0.81–0.98) | 0.89 (0.81–0.98) | 0.90 (0.82–0.99) |

| Asia-Pacific BMI classification | | Person-years | Failure | IR‡ | Crude | Model 1 | Model 2§ | Model 3¶ |
|---|---|---|---|---|---|---|---|---|
| | | | | | TR (95% CI) | aTR (95% CI) | aTR (95% CI) | aTR (95% CI) |
| **Women (n=4,963)** | | | | | | | | |
| <18.5 kg/$m^2$ | Underweight | 3,610.4 | 62 | 17.2 | 1.06 (1.00–1.12) | 1.06 (1.00–1.12) | 1.05 (0.99–1.11) | – |
| 18.5–22.9 kg/$m^2$ (reference) | Normal weight | 22,945.2 | 472 | 20.6 | 1.00 | 1.00 | 1.00 | – |
| 23.0–24.9 kg/$m^2$ | Overweight | 9,399.6 | 224 | 23.8 | 0.98 (0.95–1.02) | 0.98 (0.95–1.02) | 0.98 (0.95–1.01) | – |
| 25.0–29.9 kg/$m^2$ | Obesity class I | 8,762.1 | 258 | 29.4 | 0.95 (0.93–0.98) | 0.95 (0.93–0.98) | 0.95 (0.92–0.97) | – |
| ≥30.0 kg/$m^2$ | Obesity class II | 1,383.4 | 33 | 23.9 | 0.97 (0.91–1.04) | 0.97 (0.91–1.04) | 0.96 (0.90–1.03) | – |

Abbreviations: aTR, adjusted time ratio; BMI, body mass index; CI, confidence interval; CKD, chronic kidney disease; eGFR, estimated glomerular filtration rate; HbA1c, hemoglobin A1c; IR, incidence rate; TR, time ratio.

*Hypertension is defined as systolic blood pressure ≥130 mmHg and/or diastolic blood pressure ≥80 mmHg.

†Dyslipidemia is defined as serum low-density lipoprotein cholesterol ≥140 mg/dL, serum high-density lipoprotein cholesterol <40 mg/dL, and/or serum triglycerides ≥150 mg/dL.

‡Incidence rate is reported per 1,000 person-years.

¶Convergence errors were observed.

Multiple imputed variables: hypertension*, dyslipidemia†, self-reported alcohol intake, self-reported smoking status, and residential district.

Model 1: Adjusted for age category (34–59[reference]/60–69/70–100).

Model 2: Adjusted for the variable of Model 1, self-reported alcohol intake (non- or seldom-drinker [reference]/drinker), and self-reported smoking status (non- or ex-smoker [reference]/smoker).

Model 3: Adjusted for all variables of Model 2, hypertension* (no[reference]/yes), dyslipidemia† (no[reference]/yes), HbA1c values, and residential district (East[reference]/Tatsukawa/Central/Fudeoka/South/West/Yoshiwara/Yogita).

§A multiplicative term (BMI classification × self-reported smoking status) was added.
